# Provincial variability in congenital heart disease prevalence in Argentina, 2014–2019: A population-based analysis from national registry data

**DOI:** 10.1101/2025.09.04.25335135

**Authors:** María Clara Vita, Nicolás Fantozzi, Federico Méndez Ortiz, Ludmila Di Lullo, Romina Kokal, Simbad Peralta, Sandra Di Lalla

## Abstract

Congenital heart defects are a leading cause of neonatal morbidity and mortality worldwide. Early diagnosis is essential to enable timely treatment and improve patient outcomes; however, access to early detection and specialized care is unevenly distributed across regions in Argentina. This study describes the prevalence and timing of diagnosis of congenital heart defects and critical congenital heart defects among live births covered by the public health system nationwide.

We performed a cross-sectional, descriptive, ecological study including all live births between 2014 and 2019 with a diagnosis of congenital heart defects reported up to five years of age in the National Registry of Congenital Heart Diseases. We calculated prevalence rates and median age at case notification for each province to assess geographic disparities.

Out of 2,473,720 live births, 16,150 cases of congenital heart defects (prevalence 65.3 per 10,000 live births) and 3,700 cases of critical congenital heart defects (15.0 per 10,000) were identified. Provincial prevalence ranged widely, from 34.9 to 212.2 per 10,000 for congenital heart defects and from 10.7 to 27.3 per 10,000 for critical cases. The national median age at notification was 75 days for all congenital heart defects and 29 days for critical cases, with notable provincial differences.

These findings demonstrate significant provincial variability in both the prevalence of congenital heart defects and the age at which cases are reported to the national health registry. Strengthening early detection efforts and ensuring equitable access to specialized care are crucial to reduce morbidity and mortality associated with these conditions in Argentina and similar settings.

## INTRODUCTION

Congenital heart defects (CHD) are structural or functional abnormalities of the heart that originate during prenatal development and can be detected before or after birth. They represent the most common congenital malformation in newborns and are a leading cause of infant mortality and disability worldwide (1). In Argentina, congenital heart defects are the second leading cause of death in children under one year of age, accounting for over 10% of deaths in this age group (2).

Early diagnosis is possible through neonatal screening using pulse oximetry and prenatal diagnosis, which can identify most congenital heart defects from the 20th week of gestation (3). Advances in cardiology, cardiovascular surgery, and postoperative care have improved survival and quality of life for children with complex heart defects.

Approximately one in 500 live births is affected by critical congenital heart defects (CCHD), defined as ductus-dependent anomalies that can cause death or require invasive procedures within the first 28 days of life (4,5). These conditions pose significant clinical and public health challenges, especially in low- and middle-income countries where access to specialized care is limited and mortality rates remain high (6).

Given that nearly half of children with congenital heart defects require surgery within their first year of life and that timely diagnosis and treatment could benefit up to two-thirds of these patients, ensuring equitable access to care is essential. Barriers include limited infrastructure and human resources within the public health system to meet the national demand for diagnosis and treatment (7).

Since 2008, Argentina has implemented a National Congenital Heart Disease Program (“Programa Nacional de Cardiopatías Congénitas”) aimed at guaranteeing diagnosis and follow-up of affected patients (7). This program includes a Referral Coordination Center based in the Pediatric Hospital “Prof. Dr. Juan P. Garrahan”, which prioritizes cases based on urgency, geographic distance, and facility capacity. Each province has at least one designated referral center with a cardiologist responsible for diagnosis and reporting, facilitating timely referrals to specialized hospitals (7,8). Integration of the National Maternal and Child Health Plan has further optimized the functioning of pediatric cardiovascular surgery centers and reduced waiting times (8).

The objective of this study was to estimate the prevalence of congenital heart defects and critical congenital heart defects among live births in the public health sector from 2014 to 2019, and to analyze differences in their distribution across Argentine provinces.

## MATERIALS AND METHODS

### Study design and population

We conducted a cross-sectional, descriptive, ecological study including all live births diagnosed with congenital heart defects and registered in the National Registry of Congenital Heart Diseases (RNCC), part of the Argentine Integrated Health Information System (SISA), between January 1, 2014, and December 31, 2019. Only cases reported within the first five years of life were included. Patients of non-Argentine nationality diagnosed within the national territory were excluded.

### Definition of critical congenital heart defects

Critical congenital heart defects (CCHD) were defined as ductus-dependent structural cardiac anomalies requiring surgical or invasive intervention (surgery or cardiac catheterization) within the first 28 days of life to prevent death. Without timely intervention, mortality and survival with significant disability are extremely high (4,9). The critical defects included in this study were: aortic arch hypoplasia, aortic valve atresia, coarctation of the aorta, coronary anomaly (including ALCAPA – Anomalous Left Coronary Artery from the Pulmonary Artery), Ebstein’s anomaly, hypoplastic left heart syndrome, interruption of aortic arch, pulmonary atresia, single ventricle, transposition of the great arteries, tetralogy of Fallot, total anomalous pulmonary venous return, and truncus arteriosus.

### Data sources

- National Registry of Congenital Heart Diseases (RNCC) — SISA module (10)
- National Live Births Registry from the Ministry of Health — Directorate of Health Statistics and Information (DEIS) (11)
- M.A.P.A. platform (Monitoring for Analysis and Planning of Actions) for health workforce data including nursing, medicine, and medical specialties (Ministry of Health, 2020) (12)

### Variables

- Cumulative prevalence of CHD: calculated as the total number of registered cases during 2014–2019 divided by the total number of live births occurring in public health facilities during the same period.
- Cumulative prevalence of CCHD: calculated similarly but considering only cases classified as critical. Both prevalences are expressed per 10,000 live births and stratified by province and Buenos Aires City.
- Age at diagnosis: extracted from the RNCC based on the date of case notification.
- Sociodemographic variables: province of usual residence of the patient was included to analyze regional prevalence distribution.

### Procedure

Registered variables included diagnosis or notification date, age at notification, type of CHD, and province of residence. Prevalence estimates at the national and jurisdictional levels were calculated using the total number of cases registered relative to live births in official health establishments according to DEIS data for 2014–2019. Prevalences were calculated assuming a Poisson distribution and expressed per 10,000 live births with 95% confidence intervals.

### Methodological considerations

- Data quality control: RNCC records were validated based on completeness and internal consistency criteria. Duplicate records were excluded from analysis.
- Limitations: Prevalence estimates may be affected by underreporting in some jurisdictions, possibly due to barriers in timely notification to the surveillance system. The database includes only cases reported in public sector facilities, so results are representative only of this subsector. Public sector live births were used as the denominator to improve accuracy of estimates. Another relevant limitation is that the study period (2014–2019) predates the approval of Argentina’s Voluntary Termination of Pregnancy law in December 2020, so the potential impact of this legislation on prenatal detection and pregnancy outcomes with severe congenital heart defects could not be assessed.

### Ethical considerations

This study was conducted using secondary data from the RNCC of SISA. Data were accessed for research purposes between 1 March 2022 and 30 June 2022. Although the original dataset contained administrative variables that could potentially identify individual participants, the authors did not use or retain any such information. All analyses were conducted on de-identified data, ensuring confidentiality. The study protocol was reviewed and approved by the Ethics Committee of the Hospital General de Niños Pedro de Elizalde in Buenos Aires, in accordance with the principles of the Declaration of Helsinki (approval number: 6627).

## RESULTS

Between January 1, 2014, and December 31, 2019, a total of 2,473,720 live births were recorded in public health facilities, with 16,150 cases of CHD identified. The cumulative prevalence was 65.3 per 10,000 live births (95% CI: 64.3–66.3). The lowest provincial prevalences were observed in Jujuy (34.9; 95% CI: 30.1–40.4), Buenos Aires City (CABA) (35.2; 95% CI: 31.3–39.7), and Santa Fe (47.7; 95% CI: 44.4–51.2), while the highest prevalences were recorded in Corrientes (112.6; 95% CI: 105.6–120.1), Chubut (174.9; 95% CI: 158.3–193.1), and Tierra del Fuego (212.2; 95% CI: 183.0–246.1) (Table 1).

**Table 1.**
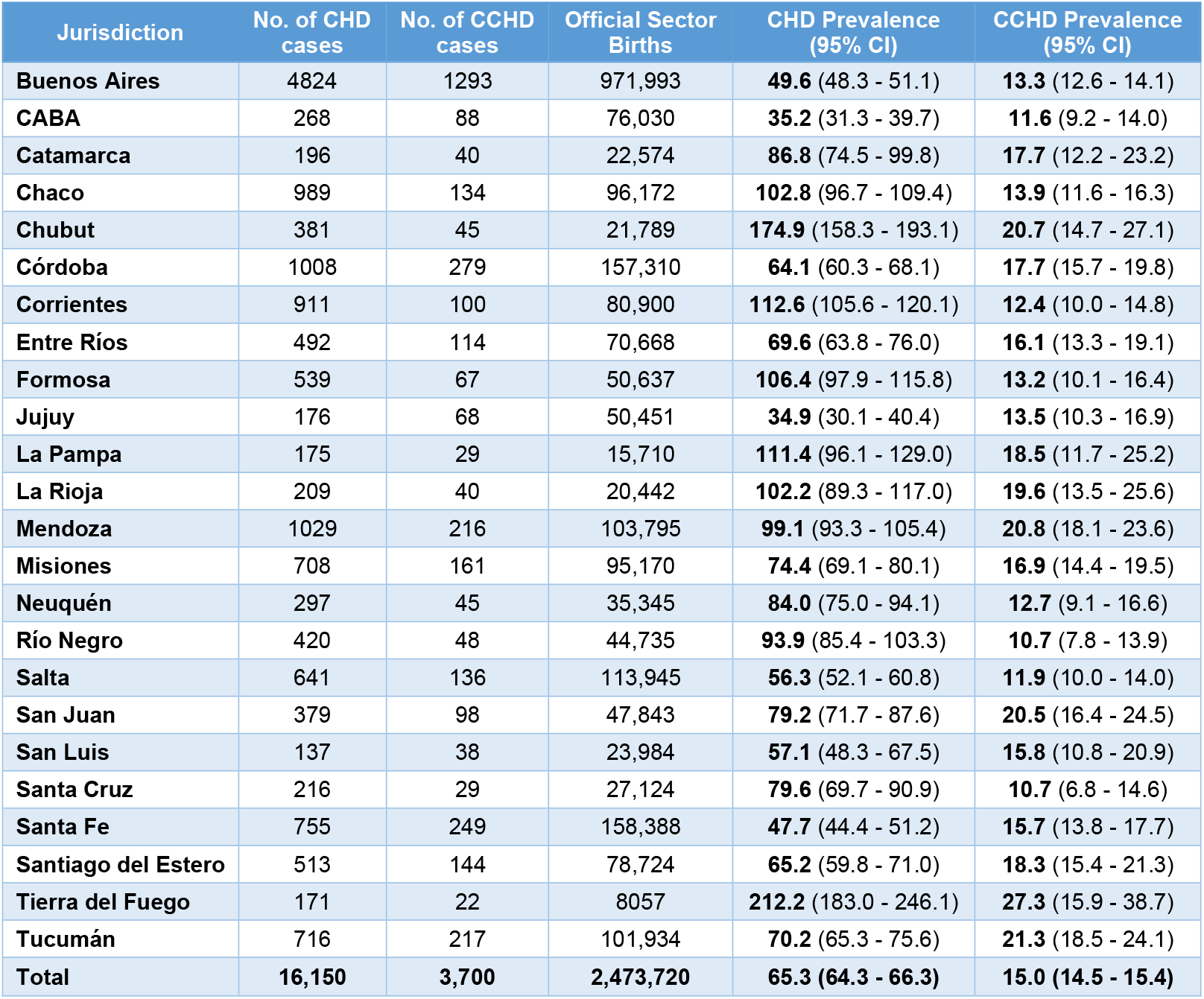
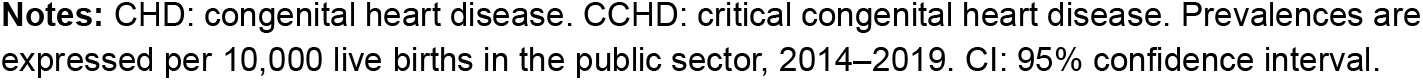
Prevalence of congenital heart disease (CHD) and critical congenital heart disease (CCHD) by jurisdiction. Argentina, 2014–2019

| Jurisdiction | No. of CHD cases | No. of CCHD cases | Official Sector Births | CHD Prevalence (95% CI) | CCHD Prevalence (95% CI) |
| --- | --- | --- | --- | --- | --- |
| Buenos Aires | 4824 | 1293 | 971,993 | **49.6** (48.3 - 51.1) | **13.3** (12.6 - 14.1) |
| CABA | 268 | 88 | 76,030 | **35.2** (31.3 - 39.7) | **11.6** (9.2 - 14.0) |
| Catamarca | 196 | 40 | 22,574 | **86.8** (74.5 - 99.8) | **17.7** (12.2 - 23.2) |
| Chaco | 989 | 134 | 96,172 | **102.8** (96.7 - 109.4) | **13.9** (11.6 - 16.3) |
| Chubut | 381 | 45 | 21,789 | **174.9** (158.3 - 193.1) | **20.7** (14.7 - 27.1) |
| Córdoba | 1008 | 279 | 157,310 | **64.1** (60.3 - 68.1) | **17.7** (15.7 - 19.8) |
| Corrientes | 911 | 100 | 80,900 | **112.6** (105.6 - 120.1) | **12.4** (10.0 - 14.8) |
| Entre Ríos | 492 | 114 | 70,668 | **69.6** (63.8 - 76.0) | **16.1** (13.3 - 19.1) |
| Formosa | 539 | 67 | 50,637 | **106.4** (97.9 - 115.8) | **13.2** (10.1 - 16.4) |
| Jujuy | 176 | 68 | 50,451 | **34.9** (30.1 - 40.4) | **13.5** (10.3 - 16.9) |
| La Pampa | 175 | 29 | 15,710 | **111.4** (96.1 - 129.0) | **18.5** (11.7 - 25.2) |
| La Rioja | 209 | 40 | 20,442 | **102.2** (89.3 - 117.0) | **19.6** (13.5 - 25.6) |
| Mendoza | 1029 | 216 | 103,795 | **99.1** (93.3 - 105.4) | **20.8** (18.1 - 23.6) |
| Misiones | 708 | 161 | 95,170 | **74.4** (69.1 - 80.1) | **16.9** (14.4 - 19.5) |
| Neuquén | 297 | 45 | 35,345 | **84.0** (75.0 - 94.1) | **12.7** (9.1 - 16.6) |
| Río Negro | 420 | 48 | 44,735 | **93.9** (85.4 - 103.3) | **10.7** (7.8 - 13.9) |
| Salta | 641 | 136 | 113,945 | **56.3** (52.1 - 60.8) | **11.9** (10.0 - 14.0) |
| San Juan | 379 | 98 | 47,843 | **79.2** (71.7 - 87.6) | **20.5** (16.4 - 24.5) |
| San Luis | 137 | 38 | 23,984 | **57.1** (48.3 - 67.5) | **15.8** (10.8 - 20.9) |
| Santa Cruz | 216 | 29 | 27,124 | **79.6** (69.7 - 90.9) | **10.7** (6.8 - 14.6) |
| Santa Fe | 755 | 249 | 158,388 | **47.7** (44.4 - 51.2) | **15.7** (13.8 - 17.7) |
| Santiago del Estero | 513 | 144 | 78,724 | **65.2** (59.8 - 71.0) | **18.3** (15.4 - 21.3) |
| Tierra del Fuego | 171 | 22 | 8057 | **212.2** (183.0 - 246.1) | **27.3** (15.9 - 38.7) |
| Tucumán | 716 | 217 | 101,934 | **70.2** (65.3 - 75.6) | **21.3** (18.5 - 24.1) |
| **Total** | **16,150** | **3,700** | **2,473,720** | **65.3** (64.3 - 66.3) | **15.0** (14.5 - 15.4) |

**Notes:** CHD: congenital heart disease. CCHD: critical congenital heart disease. Prevalences are expressed per 10,000 live births in the public sector, 2014–2019. CI: 95% confidence interval.

The national median age at the time of CHD case notification was 75 days (interquartile range [IQR]: 21–214). Provinces with the earliest notifications were San Juan (31 days; IQR: 6–127), Córdoba (39 days; IQR: 11–204), and Formosa (42 days; IQR: 9–120), whereas the highest median ages were found in Jujuy (101 days; IQR: 20–330), Santiago del Estero (121 days; IQR: 42–291), and La Rioja (147 days; IQR: 48–326) (Table 2).

**Table 2.**
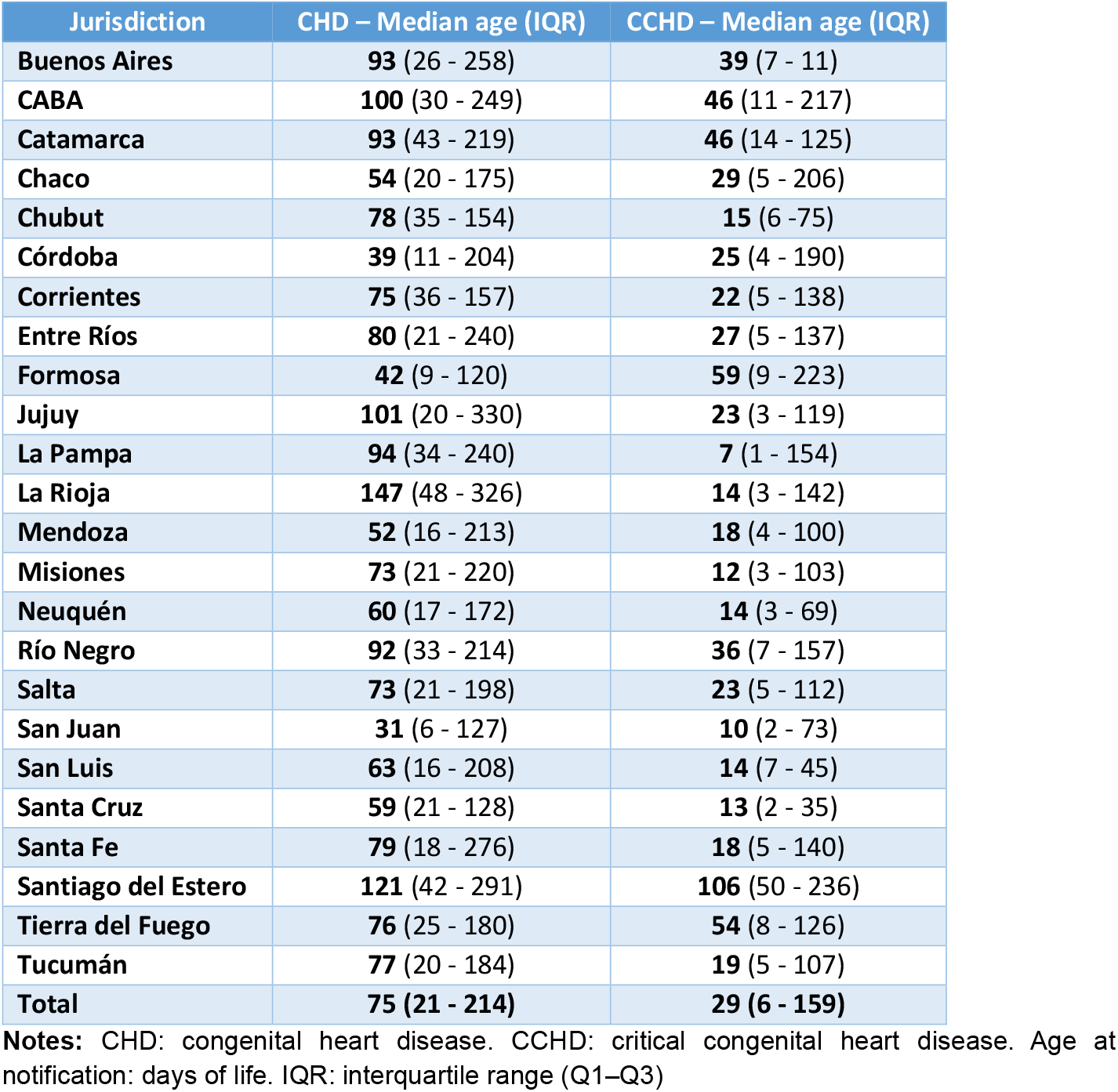
Median age at notification of congenital heart disease (CHD) and critical CHD (CCHD) by jurisdiction, Argentina, 2014–2019

| Jurisdiction | CHD – Median age (IQR) | CCHD – Median age (IQR) |
| --- | --- | --- |
| Buenos Aires | 93 (26 - 258) | 39 (7 - 11) |
| CABA | 100 (30 - 249) | 46 (11 - 217) |
| Catamarca | 93 (43 - 219) | 46 (14 - 125) |
| Chaco | 54 (20 - 175) | 29 (5 - 206) |
| Chubut | 78 (35 - 154) | 15 (6 - 75) |
| Córdoba | 39 (11 - 204) | 25 (4 - 190) |
| Corrientes | 75 (36 - 157) | 22 (5 - 138) |
| Entre Ríos | 80 (21 - 240) | 27 (5 - 137) |
| Formosa | 42 (9 - 120) | 59 (9 - 223) |
| Jujuy | 101 (20 - 330) | 23 (3 - 119) |
| La Pampa | 94 (34 - 240) | 7 (1 - 154) |
| La Rioja | 147 (48 - 326) | 14 (3 - 142) |
| Mendoza | 52 (16 - 213) | 18 (4 - 100) |
| Misiones | 73 (21 - 220) | 12 (3 - 103) |
| Neuquén | 60 (17 - 172) | 14 (3 - 69) |
| Río Negro | 92 (33 - 214) | 36 (7 - 157) |
| Salta | 73 (21 - 198) | 23 (5 - 112) |
| San Juan | 31 (6 - 127) | 10 (2 - 73) |
| San Luis | 63 (16 - 208) | 14 (7 - 45) |
| Santa Cruz | 59 (21 - 128) | 13 (2 - 35) |
| Santa Fe | 79 (18 - 276) | 18 (5 - 140) |
| Santiago del Estero | 121 (42 - 291) | 106 (50 - 236) |
| Tierra del Fuego | 76 (25 - 180) | 54 (8 - 126) |
| Tucumán | 77 (20 - 184) | 19 (5 - 107) |
| **Total** | **75 (21 - 214)** | **29 (6 - 159)** |

**Notes:** CHD: congenital heart disease. CCHD: critical congenital heart disease. Age at notification: days of life. IQR: interquartile range (Q1–Q3)

During the study period, 3,700 cases of CCHD were registered, yielding a prevalence of 15.0 per 10,000 live births (95% CI: 14.5–15.4). The lowest provincial prevalences were recorded in Río Negro (10.7; 95% CI: 7.8–13.9), Santa Cruz (10.7; 95% CI: 6.8–14.6), and CABA (11.6; 95% CI: 9.2–14.0), while the highest prevalences were observed in Mendoza (20.8; 95% CI: 18.1–23.6), Tucumán (21.3; 95% CI: 18.5–24.1), and Tierra del Fuego (27.3; 95% CI: 15.9–38.7) (Table 1).

The national median age at CCHD case notification was 29 days (IQR: 6–159). Jurisdictions with the earliest median notification ages were La Pampa (7 days; IQR: 1–154), San Juan (10 days; IQR: 2–73), and Misiones (12 days; IQR: 3– 103), while the highest median notification ages were recorded in Tierra del Fuego (54 days; IQR: 8–126), Formosa (59 days; IQR: 8.5–223), and Santiago del Estero (106 days; IQR: 50–236) (Table 2).

## DISCUSSION

This study reveals marked variability in the prevalence of CHD and CCHD across Argentine provinces. The national prevalence (65.3 per 10,000 live births) was lower than that reported in Europe (81.7 per 10,000). (13) This difference may partly reflect the inclusion in European data of live births, stillbirths, and pregnancy terminations due to fetal anomalies. (13)

At the provincial level, prevalence estimates ranged widely, with Buenos Aires City (CABA) and Jujuy falling below the national average, and provinces such as Chubut and Tierra del Fuego recording substantially higher rates. In CABA, which concentrates nearly 27% of the country’s pediatric cardiologists, the low observed prevalence likely reflects underreporting rather than access issues. (12) Since only about 33% of births in CABA occur in the public sector, cases diagnosed prenatally in private settings may not be registered if delivery occurs outside the public system. (11) Conversely, in Jujuy, limited specialist availability and diagnostic technology may hinder detection (12). Provinces with higher prevalence may reflect more intensive or efficient case notification and registration rather than a true increase in incidence. However, differences in access to services, medical resources, demographic and socioeconomic conditions, or environmental and genetic factors cannot be ruled out and warrant further research. (14) The notably high prevalence in Tierra del Fuego, both for CHD and CCHD, is particularly striking and deserves specific analysis.

The prevalence of CCHD was 15.0 per 10,000 live births, slightly higher than the 11.4 and 10.1 per 10,000 reported by Groisman et al. and Bakker et al., respectively. (15, 16) Groisman’s study analyzed data from the National Congenital Heart Disease Registry (RENAC), encompassing reports from public and private health facilities nationwide. (15) Bakker examined over 18,000 CCHD cases from 15 surveillance programs across Europe, North and South America, and Asia. (16) In that international analysis, the overall mean was 19.1 per 10,000 births, with Argentina and Slovakia showing the lowest prevalence. (16) These differences likely reflect variations in detection, coverage, and reporting systems, underscoring the need for consistent methodologies to enable valid cross-country comparisons.

Provinces such as Santa Cruz, Río Negro, and CABA showed CCHD prevalences below the national average. Given that undiagnosed CCHD can cause death within the first month of life and early detection is critical to reducing morbidity and mortality, it is essential to evaluate whether lower prevalence in these provinces relates to higher neonatal mortality. (4, 6) Limited human and diagnostic resources—especially in Santa Cruz, where there are no pediatric cardiologists according to the National Health Workforce Directorate—may contribute to incomplete case ascertainment. (12) This highlights the importance of ensuring equitable specialist coverage nationwide, alongside strengthening diagnostic capacity and surveillance systems, to improve timely case detection and reduce related morbidity and mortality.

In addition to prevalence differences, variations were observed in the timing of CHD notification across provinces, measured by the median days of life at notification. This indicator reflects the time from birth to identification and reporting of the anomaly. Nationally, the median was 75 days for all CHD cases, with San Juan, Córdoba, and Formosa having the earliest medians, and Jujuy, Santiago del Estero, and La Rioja the latest (Table 2). For CCHD, the national median time to notification was shorter, at 29 days, reflecting the greater clinical severity and detectability of these cases. Notifications occurred earliest in La Pampa, San Juan, and Misiones, while Formosa, Tierra del Fuego, and Santiago del Estero experienced the longest delays. (Table 2) These disparities likely reflect inequities in access to specialized care, healthcare workforce capacity, and the effectiveness of local surveillance systems, emphasizing the need to strengthen these aspects to improve early detection.

It is estimated that up to 25% of congenital heart defects are diagnosed late, even into adulthood. (17) Early detection of CCHD is crucial to reduce morbidity and mortality and can occur prenatally or during routine newborn examination. (4-6) However, up to 30% of newborns with CCHD are discharged undiagnosed, with mortality rates reaching 50% in this group. Thus, timely diagnosis—prenatal or postnatal—is essential to improve prognosis. (4, 6) Pulse oximetry has proven to be an effective, rapid, and non-invasive screening tool for neonatal CCHD detection. (5) Although legislation was proposed to mandate its inclusion in neonatal screening, only a few provinces have implemented it —La Pampa, Entre Ríos, and CABA—while Buenos Aires Province recommends but does not require it. This effect is evident in our data: La Pampa, which implemented the screening in 2017, showed the lowest median notification age for CCHD (7 days). In contrast, CABA (46 days) and Entre Ríos (25 days) exhibited less pronounced reductions, underscoring the importance of not only enacting legislation but also ensuring its effective implementation and monitoring to achieve timely detection.

According to the literature, CCHD represent approximately 25% of all CHD. (9) In our study, the national average was 23%, with substantial heterogeneity in the proportion of CCHD relative to total CHD across jurisdictions, as shown in Fig 1. Provinces such as Jujuy, CABA, and Santa Fe exhibited higher proportions of CCHD, whereas Corrientes, Río Negro, and Chubut showed considerably lower proportions. Jurisdictions with lower proportions may reflect challenges in detecting or reporting critical cases, early neonatal deaths without diagnosis, limited access to specialized diagnostic services, or underreporting. On the other hand, jurisdictions with high CCHD proportions may appear overrepresented due to relative underreporting of non-critical CHD cases, skewing the overall proportion. In CABA, for instance, the CCHD proportion was 33% despite one of the lowest absolute prevalences. This could reflect the concentration of high-complexity facilities and pediatric cardiology specialists, which facilitates systematic detection of critical cases, while milder or asymptomatic CHD forms may be underreported. Importantly, SISA assigns cases to the patient’s usual residence rather than place of birth. Thus, both intrauterine and postnatal referrals to high-complexity hospitals in CABA for CCHD care—when the mother or newborn resides elsewhere—are not counted as local cases. This reduces the possibility that CABA’s high CCHD proportion is solely due to referral concentration.

This study was based exclusively on live births in the public health subsector. According to current regulations, notification to SISA is mandatory for public providers, which may introduce bias, especially in provinces with a high proportion of births in the private sector. In such cases, if a child is born in the private sector but later receives public care, they may be notified as a case without the birth being included in the denominator, resulting in prevalence overestimation.

Variability in the availability of pediatric cardiologists, access to prenatal diagnostic technologies, and the implementation of strategies like pulse oximetry likely contribute to the observed heterogeneity among jurisdictions. These disparities highlight the need to review and strengthen registry systems, invest in infrastructure and training, and improve detection and care of CHD nationwide. Accurate and representative data will not only improve understanding of the distribution and magnitude of the problem but also guide effective public policies that ensure early diagnosis and timely treatment.

This study highlights marked disparities in CHD and CCHD detection across Argentine jurisdictions. Improving registry systems, expanding access to diagnostic technologies, and strengthening healthcare worker training are key steps toward earlier and more equitable diagnosis. While Argentina’s public health system offers high complexity and skilled professionals, equitable and timely access remains a challenge. Observed disparities likely reflect inequities in specialist distribution, diagnostic resources, and screening strategies, emphasizing the need to strengthen integration between health subsectors and optimize surveillance systems.

These findings have important implications for public health policy in Argentina and other middle-income countries. Targeted strategies prioritizing equity, infrastructure reinforcement, and enhanced epidemiological surveillance can improve early diagnosis and access to specialized care, reducing preventable morbidity and mortality. Robust population-based data are critical to inform interventions and policies that promote equitable child health outcomes.

## CONCLUSION

During the study period, the prevalence of CHD and CCHD in Argentina was 65.3 and 15.0 per 10,000 live births, respectively.

Substantial differences in prevalence were observed across provinces. Higher prevalence in certain jurisdictions may reflect more efficient case identification and reporting, as well as potential environmental, genetic, or sociodemographic influences that require further investigation. These findings highlight the need for ongoing research to better understand regional disparities and to guide public health strategies aimed at optimizing timely detection and management of congenital heart defects. Strengthening surveillance systems, improving access to specialized care, and ensuring equitable coverage across provinces are key steps to reduce preventable morbidity and mortality among children with congenital heart disease.

## Data Availability

The data underlying this study were derived from the Sistema Integrado de Información Sanitaria Argentino (SISA), which requires official authorization for access. The aggregated dataset used for analyses in this study is available from the corresponding author (María Clara Vita) upon reasonable request. Requests for access to the original SISA data must be directed to the appropriate authorities.

